# Knowledge, attitude, and practice of geriatric trauma risk assessment: Instrument development and validation

**DOI:** 10.1101/2023.06.23.23291834

**Authors:** Oluwaseun Adeyemi, Sanjit Konda, Corita Grudzen, Charles DiMaggio, Garrett Esper, Erin Rogers, Keith Goldfeld, Saul Blecker, Joshua Chodosh

## Abstract

**Background:** Emergency providers and nurses play pivotal roles in the initial triage and risk assessment of geriatric trauma patients. Their knowledge, attitudes, and practices of geriatric trauma risk assessment may significantly influence geriatric trauma outcomes. This study aims to develop scales that comprehensively assess emergency providers’ knowledge, attitudes, and practices of geriatric trauma triage and risk assessment.

**Methods:** We designed the knowledge (30 items), attitude (14 items), and practice (14 items) scale using the American College of Surgeons geriatric trauma management guidelines. Each of the surveys was designed using a five-point Likert scale. Content validation was performed by nine clinicians and instrument design experts. We computed Cohen’s Kappa, and item and scale content validity indices (CVIs).

**Results:** Of the 30 items in the knowledge scale, 27 were retained. The Cohen’s Kappa value ranged from 0.3 to 1.0 and the item and scale CVIs for the 27 items were each 0.90. Of the 14 items on the attitude scale, 13 were retained. The Cohen’s Kappa value ranged from 0.6 to 1.0 and the item and scale CVIs for the 13 items were each 0.94. All 14 items in the practice scale were retained. The Cohen’s Kappa value ranged from 0.6 to 1.0 and item and scale CVIs for the 14 items were each 0.86.

**Conclusion:** We present a content-validated survey instrument that can assess the knowledge, attitude, and practice of geriatric trauma risk assessment among emergency providers and nurses.

## Introduction

Geriatric trauma, defined as injuries to adults 65 years and older, poses a significant public health challenge due to older adults’ unique characteristics and vulnerabilities. In the United States (US), the population of older adults in the United States has been increasing.^1, 2^ Between 2010 and 2021, the US geriatric population grew by 38% compared to a 2% growth among the population less than 65 years.^3^ Strongly correlated with this growth is the rate of geriatric trauma, which has been increasing at an average of 4% every year.^4-6^ This rise in geriatric trauma can be attributed to various factors, including age-related physiological changes,^7-9^ frailty and co-existing comorbidities,^10-12^ and a higher risk of falls. Another challenge in managing geriatric trauma is the difficulty in accurately estimating injury severity among older adults. Traditional trauma scoring systems, such as the Injury Severity Score (ISS), do not adequately capture the complexity and impact of injuries in this population, partly due to the presence of pre-existing medical conditions, frailty, and impaired physiological reserves, which can influence the manifestation and severity of trauma-related injuries.^13-15^

Trauma triage and risk assessment are crucial components of providing optimal care for geriatric trauma patients. With the increasing population of older adults and the rising incidence of geriatric trauma, efficiently identifying and managing high-risk patients becomes crucial to avoid unnecessary hospitalizations, reduce healthcare costs, and improve overall patient outcomes.^16-18^ Also, effective trauma triage and risk assessment can help identify the severity of injuries and prioritize appropriate interventions promptly. Older adults may present with atypical symptoms or exhibit less obvious signs of trauma, making accurate diagnosis and assessment challenging.^12, 19-21^ To address challenges with geriatric trauma injury triage and risk assessment, several novel scoring tools had emerged,^13, 22-25^ and the American College of Surgeons provides regular updates on geriatric trauma management guidelines.^26^ Yet is unknown to what extent these guidelines have influenced emergency providers’ knowledge, attitude, and practice of geriatric trauma triage and risk assessment.

Emergency providers and nurses play pivotal roles in the initial assessment and management of trauma patients, including geriatric individuals. Their knowledge, attitudes, and practices significantly influence the delivery of quality care and patient outcomes. Understanding the existing knowledge, attitude, and practice gaps and identifying potential areas for improvement is essential for enhancing the care provided to geriatric trauma patients. Of the few studies that measured the knowledge, attitude, and practice of emergency providers’ geriatric trauma care,^27-30^ no study focused on US providers’ triage and post-injury risk assessments. Additionally, there are no US-validated tools to explore the knowledge, attitude, and practice of US emergency providers in geriatric trauma triage and risk assessment. This study aims to address this gap by developing scales that comprehensively assess emergency providers’ knowledge, attitudes, and practices of geriatric trauma triage and risk assessment. This scale will aid in identifying areas of improvement and inform targeted interventions to enhance the quality of care provided to geriatric trauma patients.

## Methods

### Study Design and Population

For this content validation study, we used a purposive sampling technique to select survey instrument experts to assess the content validity of the items in the knowledge, attitudes, and practice survey instruments. The selection criteria were that the instrument experts must be providers actively involved in research or non-providers with advanced degrees in epidemiological research. This study is part of validation studies aimed at developing novel tools for geriatric trauma triage and risk assessment (Institutional Review Board: s15-00371)

### Scale Development

We developed a scale to assess the knowledge, attitude, and practice of geriatric trauma triage and risk assessment. The items for scale development were adapted from the American College of Surgeons Geriatric Trauma Quality Improvement survey.^26^ The scale consisted of three independent parts: knowledge, attitude, and practice. The knowledge section included 30 items (scored from 0 to 120), the attitude section included 14 items (scored from 0 to 56), and the practice section included 14 items (scored from 0 to 56). Each item in the scale was scored on a five-point Likert scale, ranging from 1 (strongly disagree) to 5 (strongly agree). We identified certain items that required reverse coding to ensure consistency and eliminate response bias. Reverse coding was implemented for items that assessed negative perceptions or behaviors related to geriatric trauma triage and risk assessment.

### Analytical Plan

We reported the demographic and occupation characteristics of the instrument experts. We assessed the content validity of the items in the knowledge, attitude, and practice of geriatric trauma assessment by computing the content validity index (CVI).^31^ The instrument experts assessed the relevance of all items in the knowledge, attitude, and practice scales on a four-level Likert-type scale (1-irrelevant; 2-unable to assess relevance without revision; 3-relevant but needs minor alteration; 4-extremely relevant). We recoded the four-level scale into a binary scale and defined relevant items (coded as 1) as responses that are relevant or relevant with minor alterations.^31^ All other responses were coded as irrelevant (coded as 0). We computed the item content validity index (I-CVI) as the mean score of each item. Also, we computed the agreement of the experts on the relevance of each item. We generated Cohen’s kappa^32^ using the formula: 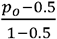 where *p*_*0*_ was the observed relevant proportion. We retained items with Cohen’s kappa value of 0.2 or greater. Additionally, we computed the scale content validity index (S-CVI) in two steps: First, we computed the proportion of experts that agree on the relevance of the items in the knowledge, attitudes, and practice scales. Then, we calculated the average of the proportions to generate the S-CVI.^31^ The survey was distributed using Research Electronic Data Capture (REDCap)^33^ and the data were analyzed with the IBM Statistical Package for Social Sciences (SPSS) version 28.^34^

## Results

### Demographic and Occupational Characteristics

Nine content and instrument experts examined the items of the knowledge, attitude, and practice of the geriatric trauma risk assessment survey (Table 1). The mean (SD) age of the experts was 34 (8.3) years and experts were predominantly male (56%) and non-Hispanic White (44%). A third of the experts were MDs and a third had doctoral degrees (PhD). The median years of practice was six years.

**Table 1:** Demographic and occupational characteristics of the content experts

| Variables | Frequency (N=9) |
| --- | --- |
| Age (Mean (SD)) | 34.0 (8.3) |
| Sex (n(%)) |  |
| Male | 5 (55.6) |
| Female | 4 (44.4) |
| Race/Ethnicity (n(%)) |  |
| Non-Hispanic White | 4 (44.4) |
| Non-Hispanic Black | 1 (11.1) |
| Multi-race | 2 (22.2) |
| Other Races | 2 (22.2) |
| Educational Qualification* (n(%)) |  |
| Master | 5 (55.6) |
| PhD | 3 (33.3) |
| M.D. | 3 (33.3) |
| Years of practice** (Median (Q1, Q3)) | 6.0 (3.0, 12.0) |

\*Multiple options, proportion exceeds 100%; Years of practice refers to both clinical and health service research

### Knowledge of Geriatric Trauma Risk Assessment

Among the 30 items in the knowledge survey, three items had Cohen’s Kappa value of less than 0.2 and these three items were removed (Table 2). The remaining 27 items had a Cohen’s Kappa value ranging from 0.3 to 1.0. The item and scale CVIs for the 27 items were each 0.90.

**Table 2:** Summary of content validity index of the items of the knowledge of geriatric trauma risk assessment survey

| Survey Items | Expert 1 | Expert 2 | Expert 3 | Expert 4 | Expert 5 | Expert 6 | Expert 7 | Expert 8 | Expert 9 | Number in Agreement | Item CVI | Kappa | Decision |
| --- | --- | --- | --- | --- | --- | --- | --- | --- | --- | --- | --- | --- | --- |
| Item 1 | 1 | 0 | 1 | 0 | 1 | 0 | 1 | 1 | 1 | 6 | 0.67 | 0.33 | Retain |
| Item 2 | 1 | 0 | 0 | 1 | 0 | 0 | 1 | 1 | 1 | 5 | 0.56 | 0.11 | Remove |
| Item 3 | 1 | 1 | 1 | 1 | 1 | 1 | 1 | 1 | 1 | 9 | 1 | 1 | Retain |
| Item 4 | 1 | 0 | 1 | 1 | 1 | 1 | 0 | 1 | 1 | 7 | 0.78 | 0.56 | Retain |
| Item 5 | 1 | 0 | 1 | 1 | 1 | 1 | 0 | 1 | 1 | 7 | 0.78 | 0.56 | Retain |
| Item 6 | 1 | 1 | 1 | 0 | 1 | 1 | 1 | 0 | 1 | 7 | 0.78 | 0.56 | Retain |
| Item 7 | 1 | 1 | 1 | 0 | 1 | 1 | 0 | 0 | 0 | 5 | 0.56 | 0.11 | Remove |
| Item 8 | 0 | 1 | 1 | 0 | 1 | 1 | 0 | 0 | 1 | 5 | 0.56 | 0.11 | Remove |
| Item 9 | 0 | 1 | 1 | 1 | 1 | 1 | 0 | 0 | 1 | 6 | 0.67 | 0.33 | Retain |
| Item 10 | 1 | 1 | 1 | 1 | 1 | 1 | 1 | 1 | 1 | 9 | 1 | 1 | Retain |
| Item 11 | 1 | 1 | 1 | 1 | 1 | 1 | 1 | 1 | 1 | 9 | 1 | 1 | Retain |
| Item 12 | 1 | 1 | 1 | 1 | 1 | 1 | 1 | 1 | 1 | 9 | 1 | 1 | Retain |
| Item 13 | 1 | 1 | 0 | 1 | 1 | 1 | 1 | 1 | 1 | 8 | 0.89 | 0.78 | Retain |
| Item 14 | 1 | 1 | 1 | 1 | 1 | 1 | 1 | 1 | 1 | 9 | 1 | 1 | Retain |
| Item 15 | 1 | 1 | 1 | 1 | 1 | 1 | 1 | 1 | 1 | 9 | 1 | 1 | Retain |
| Item 16 | 1 | 1 | 1 | 1 | 1 | 1 | 1 | 1 | 1 | 9 | 1 | 1 | Retain |
| Item 17 | 1 | 1 | 1 | 1 | 1 | 1 | 1 | 1 | 1 | 9 | 1 | 1 | Retain |
| Item 18 | 1 | 1 | 1 | 1 | 1 | 1 | 1 | 1 | 1 | 9 | 1 | 1 | Retain |
| Item 19 | 1 | 1 | 1 | 1 | 1 | 1 | 1 | 1 | 1 | 9 | 1 | 1 | Retain |
| Item 20 | 1 | 1 | 1 | 1 | 1 | 1 | 1 | 1 | 1 | 9 | 1 | 1 | Retain |
| Item 21 | 1 | 1 | 1 | 1 | 0 | 1 | 1 | 0 | 1 | 7 | 0.78 | 0.56 | Retain |
| Item 22 | 1 | 1 | 1 | 1 | 0 | 1 | 1 | 1 | 1 | 8 | 0.89 | 0.78 | Retain |
| Item 23 | 1 | 1 | 1 | 1 | 1 | 1 | 1 | 1 | 1 | 9 | 1 | 1 | Retain |
| Item 24 | 1 | 1 | 1 | 1 | 1 | 1 | 1 | 1 | 1 | 9 | 1 | 1 | Retain |
| Item 25 | 1 | 1 | 1 | 1 | 1 | 1 | 1 | 1 | 0 | 8 | 0.89 | 0.78 | Retain |
| Item 26 | 1 | 1 | 1 | 1 | 0 | 1 | 0 | 1 | 1 | 7 | 0.78 | 0.56 | Retain |
| Item 27 | 1 | 1 | 1 | 1 | 1 | 1 | 0 | 1 | 1 | 8 | 0.89 | 0.78 | Retain |
| Item 28 | 1 | 1 | 1 | 1 | 0 | 1 | 0 | 1 | 1 | 7 | 0.78 | 0.56 | Retain |
| Item 29 | 1 | 1 | 1 | 1 | 1 | 1 | 0 | 1 | 0 | 7 | 0.78 | 0.56 | Retain |
| Item 30 | 1 | 1 | 1 | 1 | 0 | 1 | 1 | 1 | 1 | 8 | 0.89 | 0.78 | Retain |
| Proportion Relevant | 0.93 | 0.87 | 0.93 | 0.87 | 0.8 | 0.93 | 0.7 | 0.83 | 0.9 | Mean CVI = 0.86 |  | Adjusted Mean Item CVI = 0.90 |  |
| Adjusted relevant proportion | 0.96 | 0.89 | 0.96 | 0.93 | 0.81 | 0.96 | 0.74 | 0.89 | 0.93 | Scale CVI= 0.86 |  | Adjusted Scale CVI = 0.90 |  |

Adjusted values are computed after removing the items with kappa values $\leq 2$ .
CVI: Content Validity Index

### Attitudes Towards Geriatric Trauma Risk Assessment

Among the 14 items in the attitude survey, one item had a Cohen’s Kappa value of less than 0.2 and this item was removed (Table 3). The remaining 13 items had a Cohen’s Kappa value ranging from 0.6 to 1.0. The item and scale CVIs for the 13 items were each 0.94.

**Table 3:**
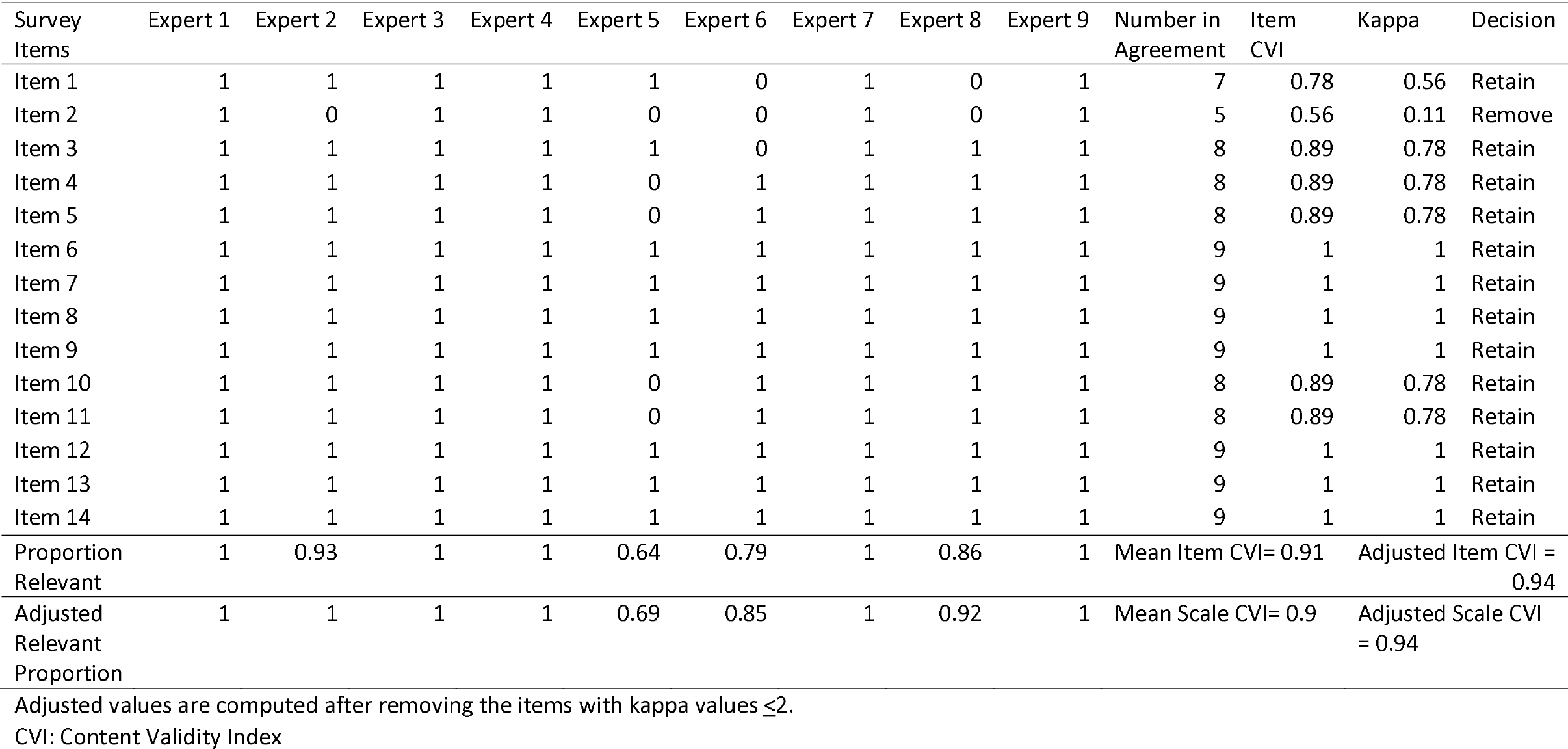
Summary of content validity index of the items in the attitudes towards geriatric trauma risk assessment survey

| Survey Items | Expert 1 | Expert 2 | Expert 3 | Expert 4 | Expert 5 | Expert 6 | Expert 7 | Expert 8 | Expert 9 | Number in Agreement | Item CVI | Kappa | Decision |
| --- | --- | --- | --- | --- | --- | --- | --- | --- | --- | --- | --- | --- | --- |
| Item 1 | 1 | 1 | 1 | 1 | 1 | 0 | 1 | 0 | 1 | 7 | 0.78 | 0.56 | Retain |
| Item 2 | 1 | 0 | 1 | 1 | 0 | 0 | 1 | 0 | 1 | 5 | 0.56 | 0.11 | Remove |
| Item 3 | 1 | 1 | 1 | 1 | 1 | 0 | 1 | 1 | 1 | 8 | 0.89 | 0.78 | Retain |
| Item 4 | 1 | 1 | 1 | 1 | 0 | 1 | 1 | 1 | 1 | 8 | 0.89 | 0.78 | Retain |
| Item 5 | 1 | 1 | 1 | 1 | 0 | 1 | 1 | 1 | 1 | 8 | 0.89 | 0.78 | Retain |
| Item 6 | 1 | 1 | 1 | 1 | 1 | 1 | 1 | 1 | 1 | 9 | 1 | 1 | Retain |
| Item 7 | 1 | 1 | 1 | 1 | 1 | 1 | 1 | 1 | 1 | 9 | 1 | 1 | Retain |
| Item 8 | 1 | 1 | 1 | 1 | 1 | 1 | 1 | 1 | 1 | 9 | 1 | 1 | Retain |
| Item 9 | 1 | 1 | 1 | 1 | 1 | 1 | 1 | 1 | 1 | 9 | 1 | 1 | Retain |
| Item 10 | 1 | 1 | 1 | 1 | 0 | 1 | 1 | 1 | 1 | 8 | 0.89 | 0.78 | Retain |
| Item 11 | 1 | 1 | 1 | 1 | 0 | 1 | 1 | 1 | 1 | 8 | 0.89 | 0.78 | Retain |
| Item 12 | 1 | 1 | 1 | 1 | 1 | 1 | 1 | 1 | 1 | 9 | 1 | 1 | Retain |
| Item 13 | 1 | 1 | 1 | 1 | 1 | 1 | 1 | 1 | 1 | 9 | 1 | 1 | Retain |
| Item 14 | 1 | 1 | 1 | 1 | 1 | 1 | 1 | 1 | 1 | 9 | 1 | 1 | Retain |
| Proportion Relevant | 1 | 0.93 | 1 | 1 | 0.64 | 0.79 | 1 | 0.86 | 1 | Mean Item CVI= 0.91 |  | Adjusted Item CVI = 0.94 |  |
| Adjusted Relevant Proportion | 1 | 1 | 1 | 1 | 0.69 | 0.85 | 1 | 0.92 | 1 | Mean Scale CVI= 0.9 |  | Adjusted Scale CVI = 0.94 |  |

Adjusted values are computed after removing the items with kappa values $\leq 2$ .
CVI: Content Validity Index

### Practice of Geriatric Trauma Risk Assessment

Among the 14 items in the attitude survey, Cohen’s Kappa value ranged from 0.6 to 1.0 (Table 4). All the items were retained. The item and scale CVIs for the 14 items were each 0.86.

**Table 3:** Summary of content validity index of the items in the practice of geriatric trauma risk assessment survey

| Survey Items | Expert 1 | Expert 2 | Expert 3 | Expert 4 | Expert 5 | Expert 6 | Expert 7 | Expert 8 | Expert 9 | Number in Agreement | Item CVI | Kappa | Decision |
| --- | --- | --- | --- | --- | --- | --- | --- | --- | --- | --- | --- | --- | --- |
| Item 1 | 1 | 1 | 1 | 1 | 1 | 0 | 1 | 1 | 1 | 8 | 0.89 | 0.78 | Retain |
| Item 2 | 1 | 1 | 0 | 1 | 0 | 0 | 1 | 1 | 1 | 6 | 0.67 | 0.33 | Retain |
| Item 3 | 1 | 1 | 1 | 1 | 1 | 0 | 1 | 1 | 1 | 8 | 0.89 | 0.78 | Retain |
| Item 4 | 1 | 1 | 1 | 1 | 0 | 1 | 1 | 1 | 1 | 8 | 0.89 | 0.78 | Retain |
| Item 5 | 1 | 1 | 1 | 1 | 1 | 1 | 1 | 1 | 1 | 9 | 1 | 1 | Retain |
| Item 6 | 1 | 1 | 1 | 1 | 0 | 1 | 1 | 1 | 1 | 8 | 0.89 | 0.78 | Retain |
| Item 7 | 1 | 1 | 1 | 1 | 1 | 1 | 1 | 1 | 1 | 9 | 1 | 1 | Retain |
| Item 8 | 1 | 1 | 1 | 1 | 0 | 1 | 1 | 1 | 1 | 8 | 0.89 | 0.78 | Retain |
| Item 9 | 1 | 1 | 1 | 1 | 0 | 1 | 1 | 1 | 1 | 8 | 0.89 | 0.78 | Retain |
| Item 10 | 1 | 1 | 1 | 1 | 0 | 1 | 0 | 1 | 1 | 7 | 0.78 | 0.56 | Retain |
| Item 11 | 1 | 1 | 1 | 1 | 0 | 1 | 0 | 1 | 1 | 7 | 0.78 | 0.56 | Retain |
| Item 12 | 1 | 1 | 1 | 1 | 1 | 1 | 0 | 1 | 1 | 8 | 0.89 | 0.78 | Retain |
| Item 13 | 1 | 1 | 1 | 1 | 0 | 1 | 0 | 1 | 1 | 7 | 0.78 | 0.56 | Retain |
| Item 14 | 1 | 1 | 1 | 1 | 0 | 1 | 0 | 1 | 1 | 7 | 0.78 | 0.56 | Retain |
| Proportion Relevant | 1 | 1 | 0.93 | 1 | 0.36 | 0.79 | 0.64 | 1 | 1 | Item CVI = 0.86<br>Scale CVI: 0.86 |  |  |  |

## Discussion

We present a content-validated instrument suitable for assessing the knowledge, attitudes, and practice of geriatric trauma risk assessment of emergency providers and nurses. With the survey instruments designed based on the American College of Surgeon’s geriatric trauma guidelines, this knowledge, attitude, and practice survey can serve as a tool to assess baseline measures, identify areas for educational intervention for geriatric trauma risk assessment, and assess the impact of interventions aimed at improving geriatric trauma triage and risk assessment.

Of the original 30 items in the knowledge survey, three were dropped due to low agreement among the experts. One of the three items that were removed assessed the last meal eaten (item 2). Similarly, the only item that was removed from the attitude survey was the last meal eaten. Meal history is relevant in assessing hypoglycemia and the risk of aspiration. In the acutely injured, it is unlikely that the response to items that assessed what was eaten and when it was eaten would affect care plans. A nasogastric tube is typically placed if there is a suspicion of risk of aspiration, even if the patient or caregivers claims the last meal eaten was eight hours.^35, 36^ Also, a bedside blood glucose check is routinely performed for geriatric trauma patients to objectively screen for hypo or hyperglycemia. ^35, 36^

The remaining two items that were removed in the knowledge survey were two of the four questions that assessed alcohol and drug abuse Excluding two of the four items that assess alcohol and drug abuse suggest a disapproval for a four-item screening questionnaire. Smith and colleagues reported that a one-item alcohol screener exhibited 82% and 79% sensitivity and specificity in detecting unhealthy alcohol use, respectively.^37^ Also, Smith and colleagues had reported that a single screening item for substance use was 100 and 74 percent sensitive and specific for drug use disorder, respectively and the single item exhibited comparable accuracy as the ten-item Drug Use Screening Test.^38^ In the ED, a single item that screens for unhealthy alcohol or drug use may, therefore, be more expedient in the risk assessment of geriatric trauma patients.

This content validation has its limitations. Our instrument and content experts include both clinicians and non-clinicians with knowledge of clinical practice. Some of the items that were considered relevant may be due to a bias toward asking such questions. This may account for the wide-ranging Cohen’s kappa scores in the knowledge survey. We used a purposive sampling design to identify the experts and the lead researcher was not blinded from knowing the experts. There is a possibility of confirmability bias.^39^ It is unlikely that the lack of blinding will differentially affect the items of the survey since these independent experts did not know who else was answering the survey. Despite these limitations, this study has its strengths. It is the first known study that developed a knowledge, attitude, and practice survey to assess geriatric trauma risk assessment by emergency providers. This survey, therefore, provides a tool to practice the evaluation and development, and assessment of interventions aimed at improving geriatric trauma risk assessment.

## Conclusion

This content validation study presents an instrument that can assess the knowledge, attitudes, and practice of geriatric trauma risk assessment by emergency providers and nurses. The knowledge, attitude, and practice scales can, therefore, be used to assess baseline characteristics as well as design and evaluate interventions aimed at improving geriatric trauma risk assessment.

## Data Availability

All data produced in the present study are available upon reasonable request to the authors

